# Observational Arthritis Foundation Internet Study: Physical Activity, Pain, and Physical Function- Study Protocol

**DOI:** 10.1101/2025.08.18.25333813

**Authors:** Sydney C. Liles, Jennifer Copson, Nurten Gizem Tore, Daniel K. White

## Abstract

**Background:** Rheumatic diseases affect approximately 54 million adults in the United States and are a leading cause of pain and disability. Although physical activity is recommended to reduce pain and improve function, rheumatic diseases encompass over 100 conditions with different clinical presentations and medical management, possibly contributing to differences in exercise response. Few studies have included diverse samples across rheumatic diseases making direct comparisons in clinical presentation and activity level difficult.

**Objective:** To evaluate the feasibility of internet recruitment, validate patient-reported diagnoses, and explore associations of physical activity with pain and function across rheumatic diseases in an online cohort of adults in the United States.

**Methods:** The Observational Arthritis foundation Internet Study (OASIS): Physical Activity, Pain, and Physical Function study is an online cross-sectional observational study of adults with rheumatic diseases living in the US. Participants will be recruited using Meta ads, Arthritis Foundation mailing lists, and the ResearchMatch database. Participants will be considered eligible if they are ≥ 18 years old, live in the US, and have been diagnosed with a rheumatic disease by a physician. All participants will provide written informed consent and HIPAA release prior to participation. Participants will self-report all rheumatic diseases they have been diagnosed with, and diagnoses will be verified using provider confirmation or electronic medical records. Self-report questionnaires will be used to assess outcomes such as physical activity, pain, physical function, mental and physical health.

**Discussion:** OASIS is the first study to comparison of physical activity, pain, and physical function across a range of rheumatic diseases in a single cohort. The results from this study could identify differences in presentation or inform tailored exercise recommendations for less common rheumatic diseases.

**Funding:** This study is funded by the Arthritis Foundation and the NIH T32HD007490. The content is solely the responsibility of the authors and does not necessarily represent the official views of the National Institutes of Health or the Arthritis Foundation.

## Introduction

One in five adults in the United States report being diagnosed with a rheumatic disease,^1,2^ accounting for more than $300 billion in direct medical costs and lost wages in 2013.^3^ Rheumatic diseases, a term which encompasses over 100 conditions including osteoarthritis (OA), rheumatoid arthritis (RA), and lupus, are a leading cause of pain and functional limitations.^1,4–7^ These symptoms interfere with performing daily activities, reduce quality of life, and lead to decreased physical activity.^1,2,8–10^ Reduced physical activity in turn increases risk of other comorbidities such as obesity, cardiovascular disease, and early mortality.^11,12^

Exercise and physical activity are strongly recommended treatments for adults with rheumatic diseases especially OA^13,14^ and RA,^15,16^ two of the most prevalent. Additionally, increased physical activity is associated with reduced risk of mortality, improved muscle strength, and improved psychological health in both the general population^11,12,17,18^ and adults with OA^13,19–21^ and RA.^15,22–25^ Previous research has also shown that increasing physical activity improves pain and function in adults with OA ^13,19–21^ and RA. ^15,22–25^ Despite these benefits, there is little research examining the effects of physical activity in other types of rheumatic diseases such as lupus, psoriatic arthritis, and gout.

Observational studies are well-suited to study such differences in treatment response and clinical presentation across rheumatic diseases. However, most studies focus on adults with OA or RA, limiting the ability to compare across other rheumatic diseases and to make evidence-based recommendations for less common diseases. Therefore, additional comparisons are needed to understand how physical activity relates to pain and function in adults with different forms of rheumatic disease. This paper outlines the protocol for the Observational Arthritis Foundation Internet Study: Physical Activity, Pain, and Function (OASIS). OASIS aims to evaluate the feasibility of internet recruitment, validate patient-reported diagnoses, and explore associations of physical activity with pain and function across rheumatic diseases.

## Methods

### Study Design

The Observational Arthritis foundation Internet Study (OASIS): Physical Activity, Pain, and Physical Function study is a completely online cross-sectional observational study focusing on adults with different forms of rheumatic disease, such as osteoarthritis (OA), gout, lupus, fibromyalgia, rheumatoid arthritis (RA), and psoriatic arthritis (PsA). This study will measure physical activity, pain, and physical function on adults with rheumatic diseases living in the United States. Study findings will be reported using the STROBE guidelines for observational studies.

### Participants

Individuals will be recruited using Meta advertisements primarily through Facebook, the Arthritis Foundation mailing list, and via the ResearchMatch participant database. ResearchMatch is a national health volunteer registry that was created by several academic institutions and supported by the U.S. National Institutes of Health as part of the Clinical Translational Science Award (CTSA) program. ResearchMatch includes volunteers who have consented to be contacted by researchers about health studies for which they may be eligible. Review and approval for this study and all procedures was obtained from University of Delaware Institutional Review Board. Participants will be included if they are at least 18 years old, live in the United States, and report having a diagnosis of a rheumatic disease provided by a physician. Participants self-report diagnoses of OA, gout, fibromyalgia, RA, psoriatic arthritis (PsA), lupus, or ‘other’ rheumatic disease. Additionally, participants must have internet access, a working email address, feel comfortable answering questions in English, and be willing to have a virtual call to verify their identity. Participants will be excluded if they fail to meet any of the inclusion criteria, have been previously enrolled, or fail the identity verification check.

### Identity Verification Check

We will employ a multi-step strategy to identify individuals who purposefully falsify information to secure payment for participation. We will utilize these strategies to check the validity of reporting from study participants during the enrollment phase. The Internet Protocol (IP) address used by the participant will be analyzed using website-based software to determine if it is located within the United States and does not have evidence of fraud or abuse. Then the informed consent form will be analyzed to ensure the printed name and signed name are consistent and match the name and signature provided on the HIPAA authorization form, if there are inconsistencies in the names, not including addition of middle initials or full versions of first name, the participant will be deemed fraudulent and excluded from the study. Additionally, if the name on the HIPAA form is not the participant’s name and/or the relationship to the participant is something other than self or equivalent (ex., Doctor, friend), then the participant will be excluded from the study. Lastly, we will examine answers to the screening form to ensure that the name provided matches the informed consent and HIPAA forms. If the participant passes all these criteria, they have passed the identity verification check and will be enrolled in the study.

### Enrollment and data collection

Participants who meet study criteria, sign the informed consent and HIPAA authorization forms, and pass the identity verification check will be enrolled. Participant demographics will then be collected, including age, sex, gender, body mass index (BMI), race, whether they were Latino, veteran status, highest level of education, employment status, and annual household income. Self-reported data on their reported rheumatic disease, physical activity, and other outcomes will also be collected.

### Disease Specific Information

Participants will self-report which rheumatic diseases they have been diagnosed with by a physician and the date of diagnosis for each. Participants will select from a list of OA, gout, fibromyalgia, RA, PsA, Lupus, or Other. If other is selected, participants are able to write in any other rheumatic disease they are diagnosed with. Participants will also be able to select multiple diagnoses. If the participant reports being diagnosed with osteoarthritis, they will be asked to report which joints were affected. National Institute for Health and Care Excellence (NICE) criteria for knee OA (≥ 45 years old, activity-related knee pain, and morning stiffness lasting < 30 minutes)^26^ will be assessed in all participants who report OA and specified the knee joint being affected. NICE Criteria have been shown to be sensitive and specific in detecting symptomatic knee OA. Participants will also be asked about previous joint injuries, joint surgeries, and past and current strategies used to manage their disease related symptoms, including options such as physical therapy, diet, corticosteroid injections, immunosuppressants, nonsteroidal anti-inflammatory medications, etc. The use of glucagon-like peptide-1 (GLP-1) agonists will also be assessed.^27,28^

## Primary Outcomes

### Physical Activity

Physical activity will be assessed using the International Physical Activity Questionnaire – Short Form (IPAQ-SF). The IPAQ-SF is a seven-item questionnaire that assesses self-reported physical activity in categories of vigorous activity, moderate activity, walking, and sedentary behavior in the form of sitting.^29–31^ Participants are asked to report the total number of minutes per day and days per week that they engage in vigorous, moderate, and walking activities. The total number of metabolic equivalent units (MET)-minutes in each category is then calculated by multiplying the total number of minutes and days in each category by the MET value for each category (8.0 MET for vigorous, 4.0 for moderate, 3.3 for walking).^32^ Participants are also classified as inactive, minimally active, and health-enhancing physical activity (HEPA) active based on the total combined MET-minutes/week calculations. ^32^ HEPA active category is defined as vigorous activity on at least three days and achieving at least 1500 MET-min/wk or 7 or more days of any activity to achieve at least 3000 MET-min/wk. ^32^ Minimally active is defined as 3 or more days of vigorous activity for at least 20 min/day, 5 or more days of moderate or walking activity of at least 30 minutes/day, or 5 or more days of any combination of activity achieving at least 600 MET-min/wk. ^32^ Inactive is defined as reporting no activity or activity insufficient to meet the criteria for the other two categories. ^32^ Sedentary behavior is assessed with one question asking participants to report the total time sitting on an average weekday. ^32^ The scoring guidelines for the IPAQ-SF will be utilized for data processing, including handling outliers, truncation of data, and minimal values for activity duration.

### Pain

The primary measure that will be used to assess pain is the Patient-Reported Outcomes Measurement Information System Pain Interference Short Form 8a (PROMIS-PI 8a). The PROMIS-PI is an 8-item questionnaire that assesses self-reported pain interference that has been validated in healthy adults and multiple clinical populations.^33–35^ Additional measures of pain will include the Visual Analog Scale (VAS) for pain and the Michigan Body Map (MBM) for chronic pain. The VAS will be collected from participants who reported a diagnosis of knee OA. This single-item questionnaire asks the participants to make a mark on a 100 mm line to indicate their pain score from 0-100 where zero represents no pain and 100 represents the worst pain imaginable.^36^ The MBM will be collected from all participants, who reported chronic pain, which is pain that is persistent or recurring pain lasting for three months or longer at the time they completed the questionnaire. Participants were asked to select on a human body map all regions where they were experiencing chronic pain. This measure has been validated in adults and is commonly used during the diagnosis of fibromyalgia.^37,38^

### Physical Function

Self-reported physical function will be measured using the Patient-Reported Outcomes Measurement Information System Physical Function Short Form 6b (PROMIS-PF 6b). The PROMIS-PF is a 6-item questionnaire that assesses self-report difficulty performing daily activities such as walking and household chores. This questionnaire has been validated in both healthy and clinical populations.^34,39–41^

## Secondary Outcomes

### Health Status

Overall health status will be measured using the PROMIS Global Health (PROMIS-GH) questionnaire. This 10-item questionnaire assesses overall health and quality of life including subscale scores for mental and physical health. Participants will also complete the Charlson Comorbidity Index (CCI) questionnaire. The CCI is a 17-item questionnaire that creates a weighted score based on a range of comorbidity conditions.

### Kinesiophobia

Kinesiophobia will be assessed using the Tampa Scale of Kinesiophobia (TSK-11). This abbreviated version of the TSK is an 11-item questionnaire that evaluates an individual’s fear-avoidance of movement, fear of work-related activities, and fear of re-injury. This questionnaire has been validated among adults with chronic pain.^42^

### Diagnosis Verification

Self-reported diagnoses will be verified via physician report or electronic medical record (EMR). Participants will specify whether they will provide EMR verification of their diagnoses or if they would prefer the research team to contact their physician for verification. Participants will then either upload a screenshot or scanned document from his/her EMR where their diagnosis is specified, or their physician will complete a provided diagnosis form template. In any instances where the specified diagnosis is unclear, the research team will adjudicate to determine whether the diagnosis can be confirmed.

## Analysis plan

We will use descriptive statistics, mean and standard deviation for continuous variables and frequency and percent for categorical variables, to summarize participant characteristics and study outcomes. To determine feasibility of recruitment, we will calculate the time to recruit 1500 participants and the cost per study completer. We will calculate percent agreement and kappa coefficients with associated 95% confidence interval to determine validity of self-reported diagnoses. We will estimate the cross-sectional association of physical activity, pain, and physical function using linear regression adjusted for potential confounders.

## Data management and Confidentiality

All data will be collected and stored in the REDCap data management system provided by the University of Delaware Center for Human Research Coordination (CHRC). These systems are password protected and require two-factor authentication to access all data.

## Ethics

This observational study received institutional review board approval from the University of Delaware. All participants must provide written informed consent and sign the Health Insurance Portability and Accountability Act (HIPAA) release form before participation. Participants have the right to withdraw at any time without penalty.

## Discussion

The OASIS study is a novel observational study of a completely online cohort of adults with rheumatic diseases. This study aims to determine reliability of patient reported diagnoses for rheumatic diseases and address the current gap in literature regarding the relationship of physical activity, pain, and function in adults with less common rheumatic diseases. Previous studies have examined the relationship between physical activity, pain, and function in individuals with OA^13,19–21^ and RA^15,22–25^, however little research on these relationships exists for lupus, psoriatic arthritis, and other rheumatic diseases which limits the ability to provide tailored activity recommendations.

### Strengths

OASIS utilizes a novel completely online recruitment strategy. This allows for recruitment of a geographically diverse sample and inclusion of individuals from rural areas, which are typically understudied. Additionally, this is the first study to include individuals with all rheumatic diseases, allowing for comparisons across rheumatic diagnoses which were not previously possible. The use of validated outcome measures strengthens the reliability of the data, and inclusion of physician verification of diagnoses increases accuracy of rheumatic disease classification.

### Limitations

Since all outcomes will be self-report questionnaires, there is a risk of recall and/or social desirability bias. OASIS is also a cross-sectional study which limits the ability to determine causal relationships between outcomes. Additionally, the online recruitment strategy may risk excluding those less likely to have access to the internet. However, data from the Pew research center shows that in 2024 96% of adults in the US report using the internet.^43^

### Implications

Results from OASIS will inform methods to conduct internet-based recruitment for future studies. Additionally, data could potentially be used to identify differences in clinical presentation and disease specific needs for treatment across rheumatic diseases. Results could also inform tailored exercise recommendations for understudied rheumatic diseases.

## Data Availability

All data produced in the present work are contained in the manuscript

